# Reasoning Over Pre-training: Evaluating LLM Performance and Augmentation in Women’s Health

**DOI:** 10.1101/2025.05.22.25328162

**Authors:** Martha Imprialou, Nikos Kaltsas, Viktoriia Oliinyk, Tom Vigrass, Joel Schwarzmann, Rachel Rosenthal, Craig Glastonbury, Chris Wigley, Matt Gillam, Nikita Kanani, Pras Supramaniam, Ingrid Granne, Cecilia M. Lindgren

## Abstract

Recent advances in large language models (LLMs) show promise in clinical applications, but their performance in women’s health remains underexamined ^1^. We evaluated LLMs on 2,337 questions from obstetrics and gynaecology, including 1,392 from the Royal College of Obstetricians and Gynaecologists Part 2 examination (MRCOG Part 2) ^2^, a UK-based test of advanced clinical decision-making, and 945 from MedQA^3^, a dataset derived from the United States Medical Licensing Examination (USMLE). The best-performing model—OpenAI’s o1-preview^4^ enhanced with retrieval-augmented generation (RAG)^5,6^—achieved 72.00% accuracy on MRCOG Part 2 and 92.30% on MedQA, exceeding prior benchmarks by 21.6%^1^. General-purpose reasoning models outperformed domain-specific fine-tuned models such as MED-LM^7^. We also analyse performance by clinical subdomain and discover lower accuracy in areas like fetal medicine and postpartum care. These findings highlight the importance of reasoning capabilities over domain-specific fine-tuning and demonstrate the value of augmentation methods like RAG for improving accuracy and interpretability^8^.

## Introduction

Large Language Models (LLMs) have demonstrated significant potential in medical applications, indicating their promise as tools for patient health coaching and clinical decision support^9–12^. However, it remains unclear how LLMs perform in women’s heath, where documented gender disparities and biases in clinical settings pose unique scientific challenges^13,14 15^. A dedicated evaluation effort focused on women’s health is therefore warranted.

Medical licensing examinations with single-answer multiple-choice questions provide unambiguous assessment of LLM medical knowledge. For obstetrics and gynaecology, the Membership of the Royal College of Obstetricians and Gynaecologists (MRCOG) Part 1 and Part 2 examinations offer a rigorous UK-based professional qualification standard ^16^. The MRCOG Part 1 examination assesses foundational obstetric and gynaecological knowledge and is typically undertaken during early training. LLMs have demonstrated notable proficiency on this assessment—GPT-4o achieved 72.2% accuracy. However, its performance declined to 50.4% on the more clinically oriented MRCOG Part 2^1^.

The MRCOG Part 2 examination evaluates advanced clinical reasoning, problem-solving, and knowledge application, typically undertaken during later specialty training stages after completion of MRCOG Part 1 and several years of clinical experience. This challenging assessment reports international candidate pass rates ranging from 8.5% to 65.9%^17^. The examination contains two question types: Single Best Answers (SBAs) and Extended Matching Questions (EMQs). Candidates must demonstrate competence across diverse obstetric and gynaecological scenarios, reflecting the breadth of clinical practice and decision-making skills required. A typical exam comprises 100 questions—50 SBAs (40% of final grade) and 50 EMQs (60%) ^2^. SBA questions present brief clinical vignettes with multiple options requiring selection of the single most appropriate answer, while EMQs group multiple vignettes under common themes with shared answer lists, testing more nuanced clinical reasoning through best-match selection. Previously, GPT-4o achieved only 45% accuracy on EMQs compared to 54% on SBAs^1^.

In this study, we expand on previous knowledge from MRCOG Part 1^1^, and focus on LLM performance on MRCOG Part 2 by conducting a comprehensive evaluation of state-of-the-art LLMs using a set of 1392 questions from that exam, 706 Single Best Answer (SBA) questions and 686 Extended Matching Questions (EMQs) to assess whether state-of-the-art LLMs are better at applying advanced medical knowledge and reasoning.

In addition, we have evaluated LLMs using 945 women’s health questions selected from the MedQA dataset, consisting of questions from the United States Medical Licensing Examination (USMLE). We used OpenAI GPT-4o to filter the broader MedQA dataset for questions with female patient scenarios and including terminology specific to women’s physiology. All selected questions were manually validated for clinical relevance. (**Materials and Methods**). The MedQA dataset is commonly used for evaluation of LLMs^9–12^, and its questions are at a more foundational level compared to the MRCOG, as the USMLE is taken in the early stages of undergraduate medical training.

Our evaluation framework encompassed several model variants: (1) general-purpose LLMs trained on diverse internet texts (including multiple versions of OpenAI’s GPT^4^, Llama^18^, Claude^19^, Gemini^20^, and DeepSeek^21^), (2) domain-specialized models such as MEDLM^7,22^ and Med Llama3^23^ that have been fine-tuned on curated clinical datasets, and (3) augmented models using Retrieval-Augmented Generation (RAG)^5,6^, where prompts are enriched with context derived from embeddings of a women’s health knowledge base comprising 600 documents, curated with documents from the MRCOG Part 2 reading list^24^ as well as UK national guidelines. We assessed performance across these variants by reporting overall accuracy in MRCOG and MedQA datasets, accuracy stratified by MRCOG question type (SBAs, EMQs) and quantified uncertainty using bootstrapped 95% confidence intervals.

By integrating rigorous evaluation across multiple datasets and model architectures, this study aims to determine whether enhancements in clinical reasoning and contextual augmentation can close the performance gap in women’s health medical undergraduate and postgraduate examinations. Our results provide novel insights into the relative strengths of general-purpose versus specialized LLMs and emphasize the potential of RAG techniques to improve accuracy and interpretability in this challenging domain.

## Results

### LLMs performance in the MRCOG Part 2 and MedQA datasets

To calculate overall MRCOG Part 2 accuracy, we simulated the exam format through 5000 bootstrapping iterations of 100 questions (50 SBAs and 50 EMQs, sampled randomly), with SBAs weighted at 40% and EMQs at 60% of the total score^2^. Similarly, for MedQA we bootstrapped sets of 100 women’s health questions using the same approach and giving equal weight to all questions – note that our benchmark does not simulate the USMLE, as that involves questions beyond women’s health. (**Figure 1a,b**).

**Figure 1a:**
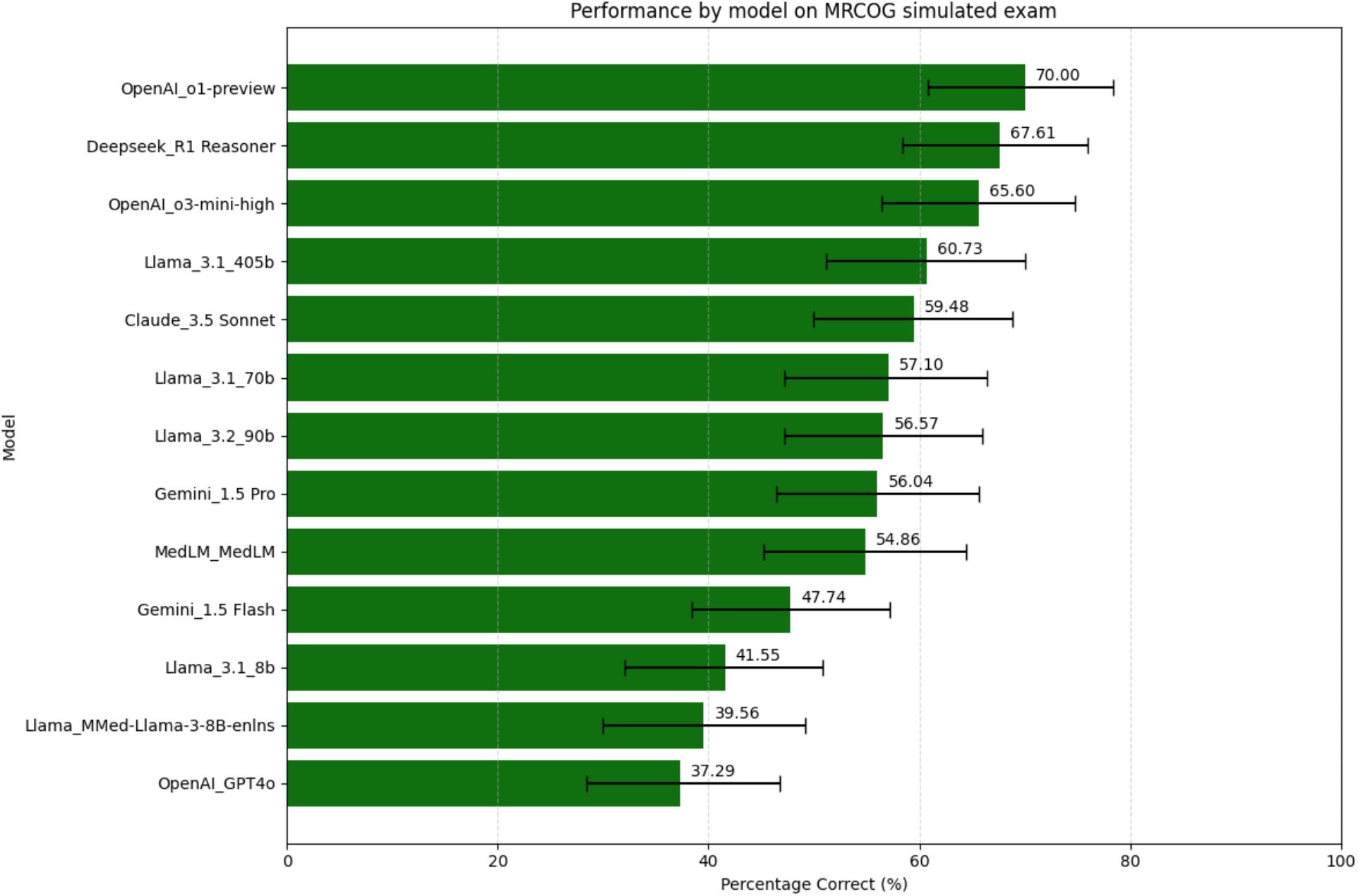
Base model performance in simulated MRCOG part 2 exam: Outlines average accuracy [%] of publicly available LLMs in answering MRCOG Part 2 questions, and 95% confidence interval from bootstrapping iterations. Each model is provided 5000 exams, each consisting of 50 SBAs and 50 EMQs randomly selected from the database of questions. The score is weighted to 60% EMQ and 40% SBA to yield an overall percentage of correct responses over 500 iterations.

**Figure 1b.**
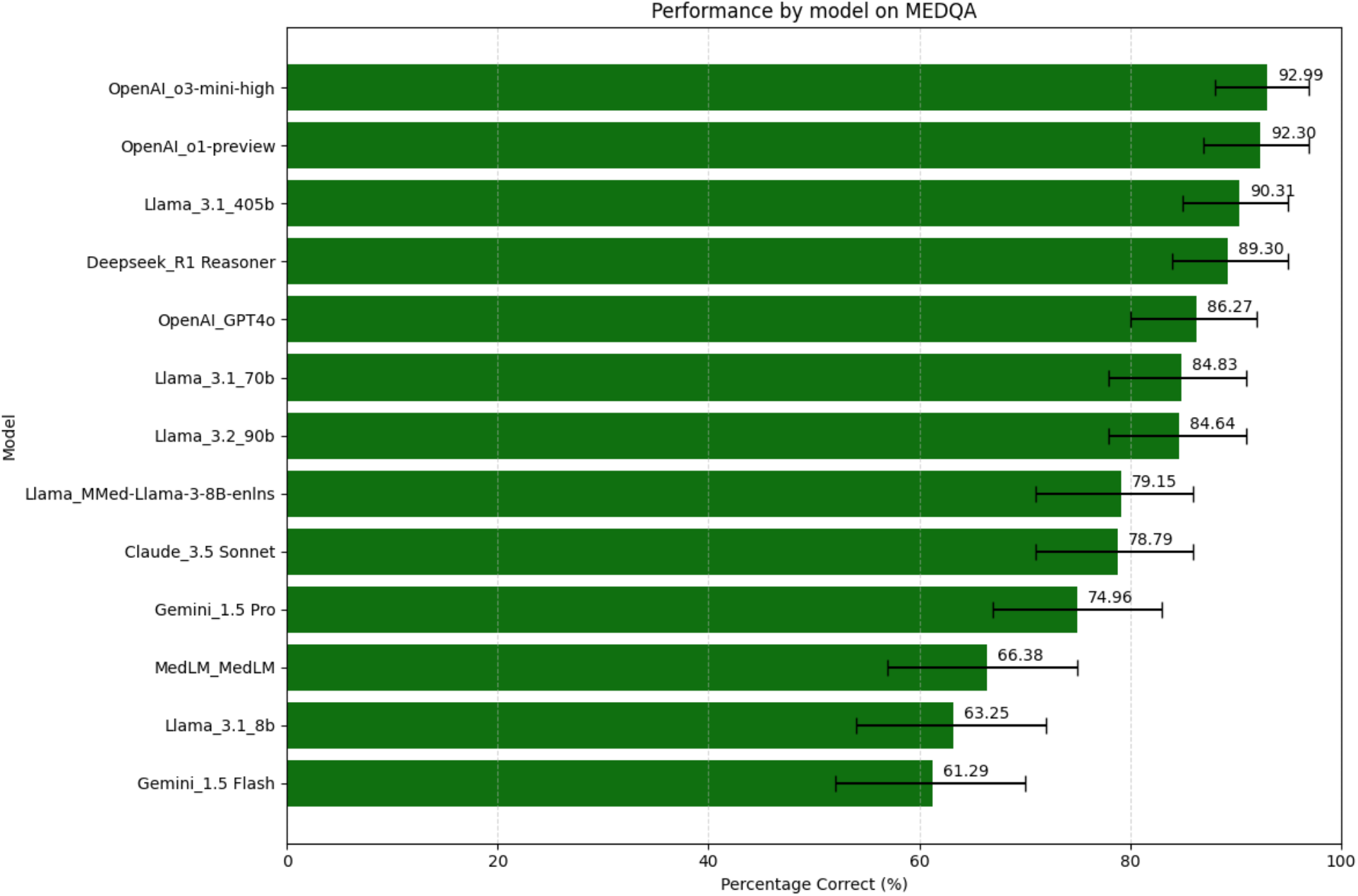
Base model performance in MedQA women’s health questions: Outlines average accuracy % of publicly available LLMs in answering MedQA questions on women’s health, and 95% confidence interval from bootstrapping iterations. Each model is provided 5000 sets of 100 questions.

Among base LLMs, OpenAI o1-preview achieves the highest average accuracy (70.00%, CI 60.80-78.40%, on MRCOG Part 2, 92.30%, CI 87-97%, on MedQA), while DeepSeek-R1, o3-mini, and Llama-3.1-405b all exceed 60% average accuracy on MRCOG Part 2 and 89% on MedQA. Domain-specific LLMs, which have been fine-tuned by pre-training with specialised medical corpora, demonstrated lower accuracy than general models, with MedLM achieving 54.86%, CI 45.2-64.4%, on MRCOG Part 2 and 66.38%, CI 57-75%, on MedQA and MMed-Llama achieving 39.60%, CI 30.0-49.2%, on MRCOG Part 2 and 79.15%, CI 71-86%, on MedQA. One possible explanation is model size: Mmed-Llama, with 8 billion parameters, was outperformed by the larger 70b and 405b parameter Llama models of the same family; indicating that very large models acquire substantial general knowledge that may allow them to address specialised questions better than fine-tuned models. The number of parameters for MedLM are not publicly known, so other aspects of algorithm design could be at play.

### RAG-enhanced LLMs performance in the MRCOG Part 2 and MedQA datasets

We then sought to test whether relevant context in prompting can increase performance, so we optimised with a RAG prompting strategy by retrieving relevant context from a medical knowledge base of 600 clinical guideline and educational documents. We performed this on a subset of the base models considered i.e. the top-performing models that had achieved a performance of >60% in MRCOG Part 2 i.e. o1-preview, DeepSeek-R1, o3-mini, Llama 3.1 405b – we also included Claude 3.5-Sonnet, which came close with 59.33%. To understand if RAG has a greater impact on previous generation models or models with smaller parameter sets, we also applied RAG on GPT-4o and Llama 3.1-8b – two of the lowest performing models in our evaluation.

RAG-enhanced models had increased accuracy compared to base models in MRCOG Part 2 (**Figure 2a**). RAG-enhanced GPT-o1 achieves the best performance of all models in this evaluation with average accuracy 72.00% (+2%).

**Figure 2.**
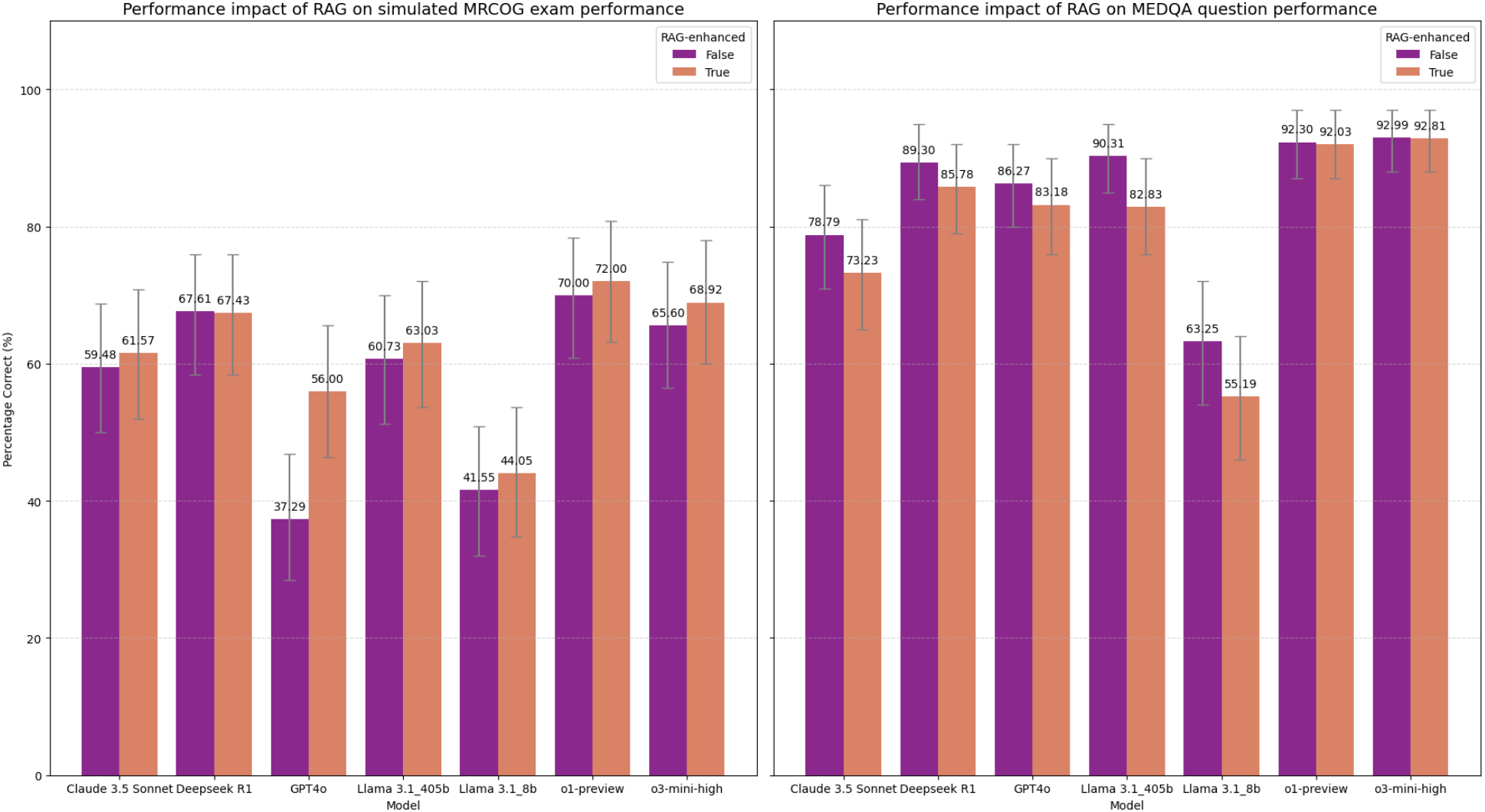
Comparison of average accuracy of RAG-enhanced LLMs vs base-LLMs across MRCOG Part 2 and MedQA. The purple bars show the performance of base LLMs when prompted only with an exam question or scenario, while the salmon bars show performance of the same models, prompted with the exam question and additional context retrieved from a medical knowledge base. Results generated via bootstrapped iterations of MRCOG Part 2 and MedQA simulations, as in **Figure 1a & 1b**.

Overall, our MRCOG Part 2 evaluation showed that RAG consistently improved performance across all models tested, including those with advanced reasoning capabilities and very large parameter counts, such as o1-preview and Llama 3.1-405b. This indicates that providing relevant contextual information within prompts can enhance the performance of models irrespective of their size or capabilities. The amount of RAG-enhanced improvement differs in each model, with lower performing base models benefiting greatly: GPT-4o improves by 1.5-fold in MRCOG (base GPT-4o average accuracy 37.29% vs 56.00% for RAG-enhanced version), while top-performing models seeing smaller gains. Additionally, considering the observation from the previous section—that models fine-tuned specifically on specialized documents achieved relatively lower overall performance—it appears that contextual prompting via RAG may be a more effective strategy than specialized pre-training alone.

In contrast, we observe that RAG-enhanced models achieve smaller performance gains in MedQA question answering, particularly for models which are already high performing where the effect is not statistically meaningful. For example, the best-performing model in MedQA, o3-mini sees a fall of 0.18% in accuracy after RAG prompting (92.99% base vs 92.81% RAG-enhanced average accuracy. 95% CI ranges are identical at 88-97%). This could be explained by the nature of the questions, which test more foundational medical knowledge and so the underlying training data for the large models trained with large volumes of data from the internet may have already ‘learned’ the domain sufficiently. Another possibility is that questions and answers from MedQA, which is frequently used in LLM benchmarking, may have been part of the training set of these models.

### LLMs performance by MRCOG question type

To explain the performance gains in the MRCOG Part 2 in the latest models against earlier studies, we compare the performance of models in specific question banks i.e. in the MRCOG SBAs and MRCOG EMQs. Previously^1^, a considerable difference of 9% in accuracy was observed between SBAs and EMQs (54% vs 45%) for GPT-4o; this difference could be attributed to the nature of the questions, as SBAs test textbook knowledge while EMQs assess clinical reasoning skills. Moreover SBAs, require choosing an answer out of 5 options, whereas EMQs offer up to 18 options. For GPT-4o, our study replicates this finding with an 8.3% difference in average accuracy for SBAs and EMQs (42.20% vs 33.89%) – on average GPT-4o performance is lower than what was previously reported^1^, which could be explained by differences in the evaluation set, as well as in changes in the deployed model by OpenAI which may have occurred since.

Crucially, our evaluation demonstrates that the previously observed performance gap between SBAs and EMQs has been effectively closed by the latest generation of models.

Models with advanced reasoning capabilities, such as OpenAI o1-preview, DeepSeek-R1, and OpenAI o3-mini-high, showed minimal differences in their performance on SBAs versus EMQs, suggesting that advanced reasoning may be particularly beneficial in accurately addressing EMQs. For example, the best-performing base model, OpenAI o1-preview, exhibits a performance difference of only 2.36% between question types, with a mean accuracy of 71.42% on SBAs (95% CI 50–90%) and 69.06% on EMQs (95% CI 50-90%). The addition of RAG further enhances performance across both question formats; notably, the RAG-enhanced OpenAI o1-preview achieves a mean accuracy of 73.31% in SBAs (95% CI 55-90%) and 70.98% in EMQs (95% CI 50-90%).

Detailed results for base models are in **Figure 3** and for RAG-enhanced models in **Figure 4**. The performance of all models in the evaluation across question type is in **Table S1** in the **Supplementary Tables**.

**Figure 3.**
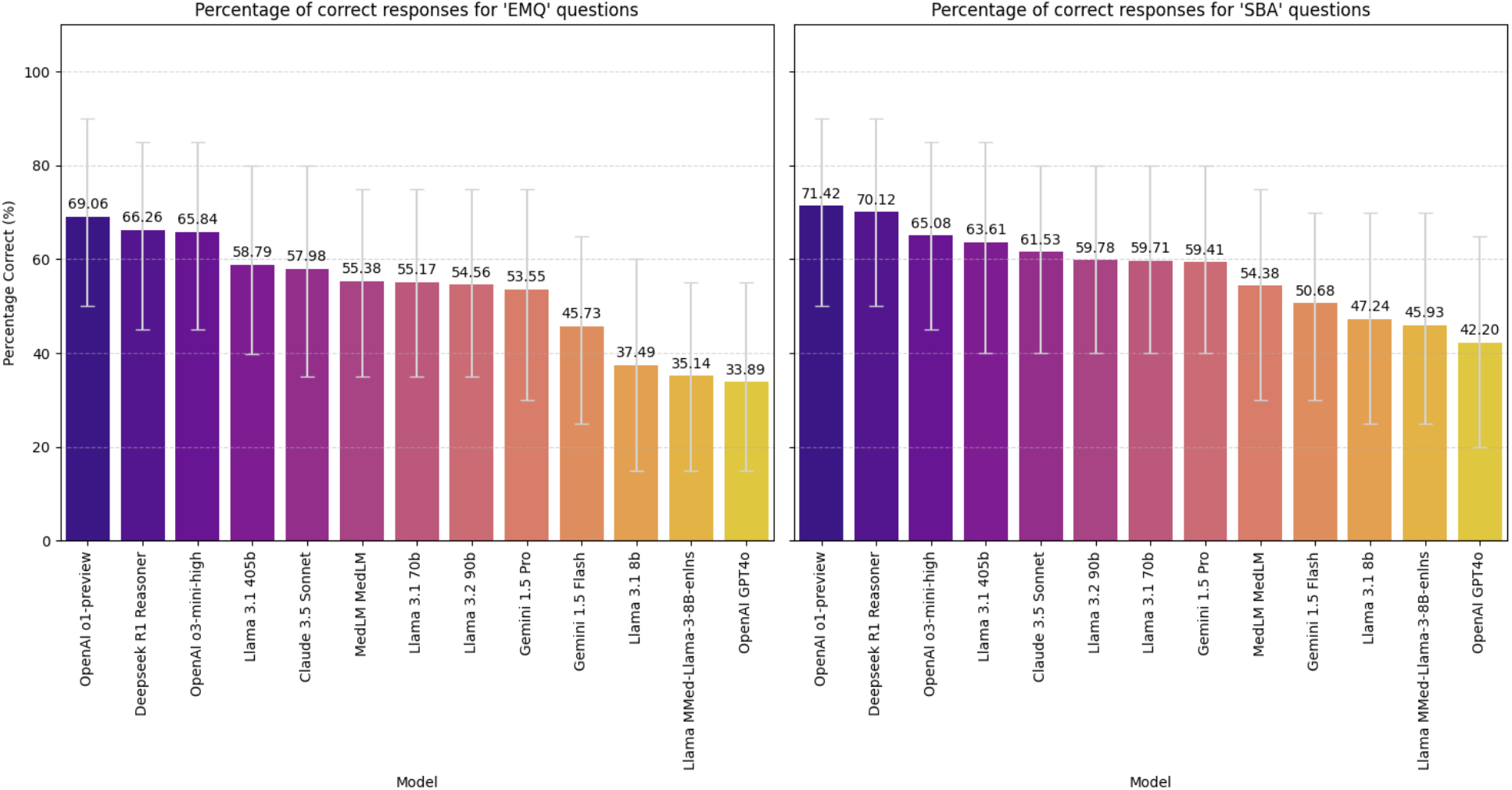
Performance of different models, broken into the different categories of questions: SBAs & EMQs: The models were provided a non-repeating random sample of 20 questions, relating to a single format of question. Average percentage of correct responses across the 20 questions is reported here over 5000 iterations of bootstrapping.

**Figure 4.**
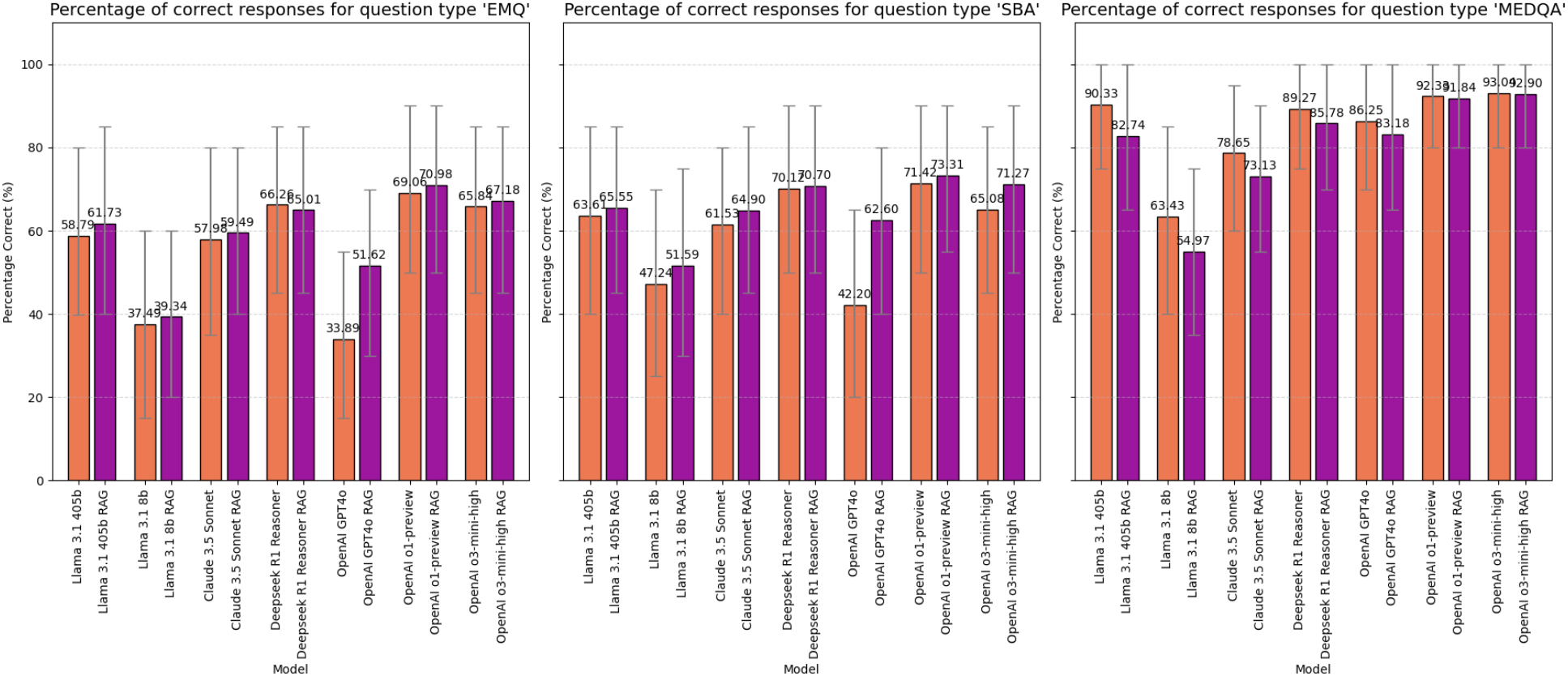
A comparison of models with and without RAG, when provided only questions of one format, i.e. SBA or EMQ. The format of the testing is equivalent to Figure 3, but these charts highlight those models where we have performed the evaluation twice, with and without RAG for comparison.

### Performance of LLMs in MRCOG Part 2 by clinical subdomains

To further understand the differences in performance of models in the MRCOG Part 2 and evaluate their usage in specific clinical scenarios we segmented the sections in clinical subdomains. The subdomains were derived from the source materials, as some of the revision books for the MRCOG Part 2 commonly group questions into specific domains. We then used Claude 3.5, Deepseek R1-reasoner and Gemini 1.5 Flash to group any unclassified questions into the same subdomains via a voting system and manually reviewed the questions and re-classified some of the questions where LLMs were unable to infer a category. Some conceptually similar categories were grouped together, to ensure each subdomain had a sufficiently large sample size to draw inference. **Table 1** summarises the subdomains we extracted and the number of MRCOG Part 2 questions in each one. Note that this analysis was performed only on the MRCOG Part 2 questions, as MedQA questions are not usually grouped in subdomains and span a wider range of topics.

**Table 1.** The question categories into which MRCOG Part 2 exam questions were classified and the number of questions present in each.

| Category | Number of Questions |
| --- | --- |
| Antenatal and Early Pregnancy Care | 237 |
| Clinical Governance, Research and Teaching | 93 |
| Fetal Medicine, Labour, Delivery, and Postpartum Care | 285 |
| Gynaecological Problems and Oncology | 274 |
| Labour, Delivery, and Postpartum Care | 228 |
| Maternal Medicine and Medical Disorders in Pregnancy | 186 |
| Sexual and Reproductive Health | 88 |
| Subfertility and Endocrinology | 138 |
| Surgical Skills and Procedures | 91 |

The results of this evaluation for the best performing general model, base and RAG-enhanced OpenAI o1-preview) is presented **Figure 5** alongside the performance of the two specialised medical models (MedLM, MMed-Llama). The performance of all models in the evaluation across subdomains can be found in **Table S2** in the **Supplementary Tables**.

**Figure 5:**
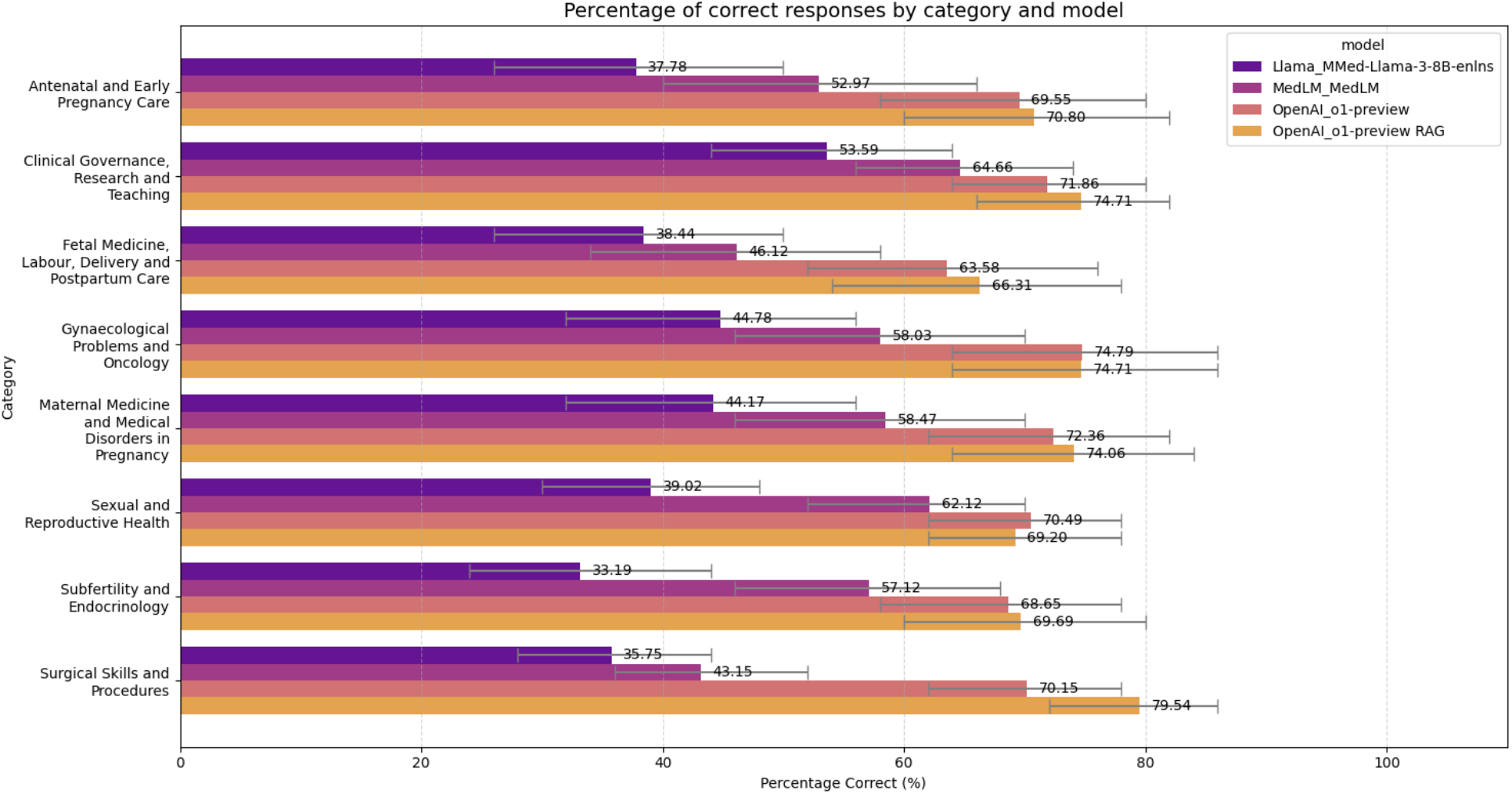
Performance of specialised, chain of thought and RAG models broken out by category of MRCOG part 2 question.

The breakdown of performance by subdomain reveals variability in the accuracy of models by question category. Overall, “Fetal Medicine, Labour, Delivery & Postpartum Care” appears as the lowest performing category on average with an average accuracy of 52.43% across all models. Clinical Governance, Research and Teaching however saw the highest performance, averaging 65.44% across all models. At the same time, we observe that the relative ranking of model performance is for the most part consistent across the different subdomains, i.e. the best performing models perform better across all sub-domains.

We observe that the specialised models (MedLM, MMed-Llama), have relatively high fluctuations in performance, with “Surgical Skills & Procedures” and “Fetal Medicine, Labour, Delivery and Postpartum Care” emerging as the worst categories, where both models score under 46% accuracy – a finding which suggests they may not be a good fit for applications that relate to those areas. In contrast the best-performing model (RAG-enhanced OpenAI o1-preview) achieves a 79.5% average accuracy (+36.7%) for “Surgical Skills and Procedures” with RAG providing a +9% in accuracy gains compared to the base model, and a 66.4% (+20.1%) average accuracy for “Fetal Medicine, Labour, Delivery and Postpartum care”.

Overall, the general-purpose OpenAI o1-preview & RAG enhanced OpenAI o1-preview, have more consistent performance across clinical subdomains in this analysis, indicating that they could perform adequately in a wide range of clinical scenarios.

The models in figure 5 were provided a miniaturised simulated exam format of 25 SBAs and 25 EMQs, non-repeating, randomly sampled and evenly weighted from a single category of medical training. The reduced size was in response to limitations in question volumes for some categories. Average percentage of correct responses across the 50 questions without differential SBA/EMQ weighting is reported here over 5000 iterations of bootstrapping.

## Discussion

The benchmarking exercise has yielded novel insights on the performance of LLMs within the underexplored field of women’s health. Utilising the largest evaluation dataset to date, we conducted comprehensive comparisons between state-of-the-art general-purpose models and their earlier iterations, as well as between specialised (fine-tuned) medical models. Our analysis incorporated a retrieval-augmented generation (RAG) approach and stratified performance by question type and by clinical subdomain.

The performance of the best performing model in our study (RAG-enhanced o1-preview) is 21.6% higher in the MRCOG Part 2 than the previous benchmark using GPT-4o^1^, indicating that state of the art models have improved considerably in medical knowledge in women’s health, with the RAG prompting strategy offering a further boost in performance but also allowing for interpretability of their reasoning.

Several base models achieved a high performance that exceeds 60% for the MRCOG Part 2 examination (Llama 3.1, Deepseek R1-Reasoner, and OpenAI o1-preview & OpenAI o3-mini-high). For example, OpenAI o1-preview consistently surpassed a 70% performance benchmark on MRCOG Part 2, while models with advanced reasoning capabilities, such as Deepseek-r1 and OpenAI o3-mini-high, also excelled in handling the nuanced EMQs designed to assess clinical decision-making and judgement. In contrast, earlier iterations (e.g., GPT-4o) exhibited considerably lower performance on these EMQs, a finding that aligns with previous evaluations^1^.

Our results indicate that RAG improves performance when it comes to EMQ and SBA type questions, on average by 10.23% for EMQ and 11.29% for SBA questions. Non-reasoning models (Llama 3.1-8b & 405b, Claude 3.5 Sonnet, GPT-4o) see a greater benefit, by 16.52% (SBA) and 16.21% (EMQ) on average, indicating that RAG can help bridge the gap in performance to reasoning models that utilise a chain-of-thought architecture (o1-preview, o3-mini, DeepSeek R1). Reasoning models saw a smaller but noticeable benefit, 4.33% (SBA) and 2.25% (EMQ), when enhanced with RAG. Consequently, investing further in RAG strategies may provide further performance gains; for example, by incrementing the knowledge base with additional content or by improving the retrieval algorithm with methods like GraphRAG^25,26^.

Additionally, we observe that model size was more predictive of performance across all our experimental setups, rather than specialised training. Specifically, in the Llama 3 family of models, MMed-Llama which is an 8 billion parameter model performed worse than the generic 70b model of the same family, which in turn was outperformed by the biggest 405 billion parameter Llama 3. Overall, our evaluation shows that general-purpose large language models outperform specialised, fine-tuned medical models in addressing women’s health queries. This suggests that the extensive pre-training on diverse datasets enables these models to manage specialised Q&A tasks on women’s health more effectively than additional fine-tuning with specialised datasets; a finding that has been reported in other domains^27–29^.

While OpenAI o1-preview demonstrates the highest performance among the evaluated models it is also significantly more expensive than the alternatives (**Figure 6**). As shown, smaller models tend to be more cost-effective and offer faster inference times but at the expense of performance. This highlights a critical trade-off in model selection, where performance, cost, and inference speed must be carefully balanced depending on the application requirements.

**Figure 6.**
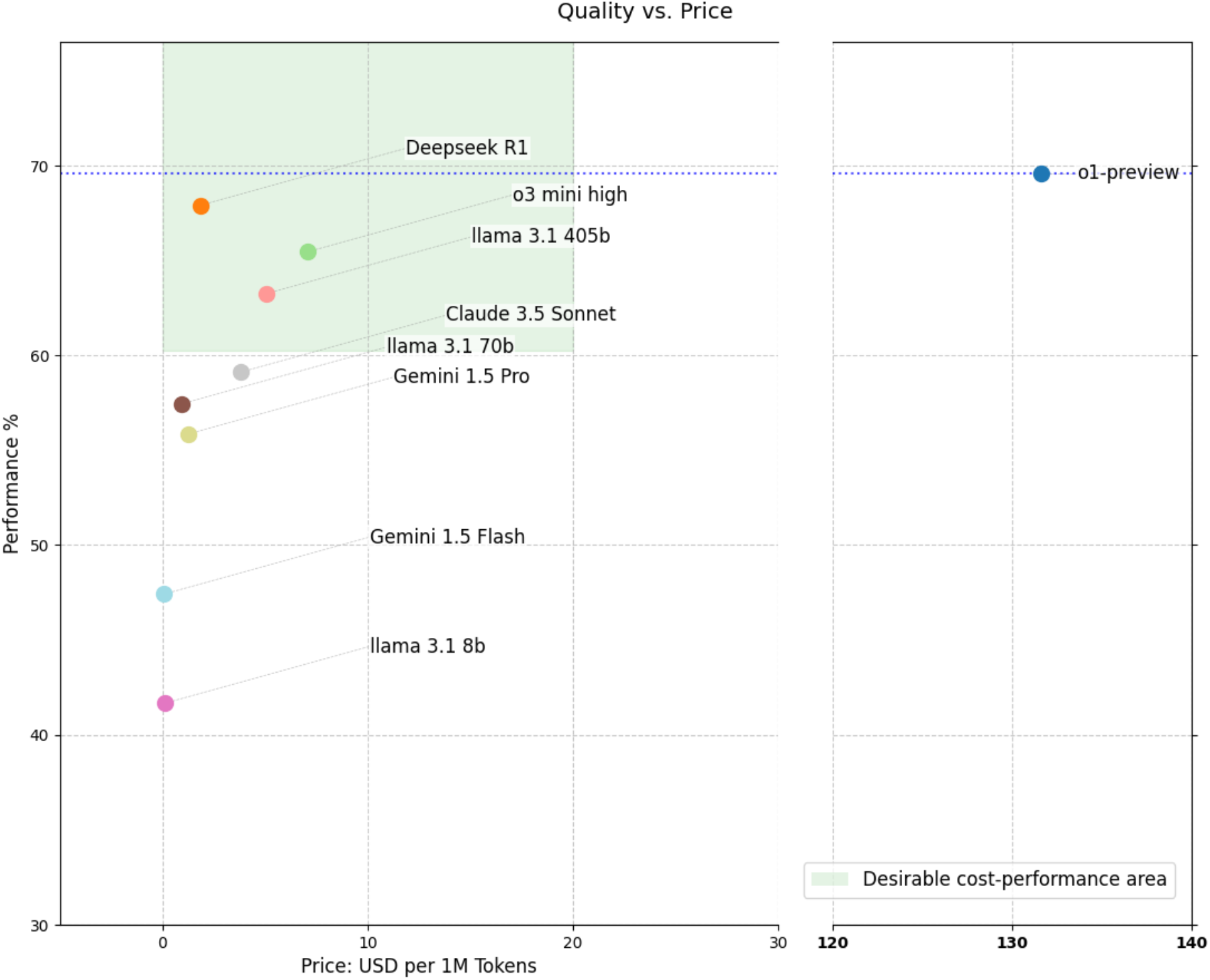
Q&A performance accuracy vs cost for the models included in the benchmark. The y-axis shows average Q&A accuracy in MRCOG Part 2, and the x-axis shows the USD cost of each model per 1M Tokens.

Finally, we have seen that while LLMs perform robustly on general clinical questions in women’s health, their accuracy can vary across specific clinical subdomains. The subdomain “Fetal Medicine, Labour, Delivery and Postpartum care” was the overall most difficult category for most of the models we tested, though there was variability in the subdomains that were considered challenging for each model – this variability may indicate gaps in the clinical knowledge represented in their training datasets. Combining multiple LLMs may help address performance issues in certain clinical subdomains.

However, our analysis highlights that the best performing model, RAG-enhanced OpenAI o1-preview, as well as the corresponding base model, had relatively consistent performance across all subdomains; a finding that strengthens the case for its utilisation in real-world clinical applications.

In summary, in this study we demonstrated that chain-of-thought reasoning LLM models introduced in early 2025, achieve robust performance on a wide range of clinical questions in women’s health, and significantly outperform earlier LLM model versions. We also found that using RAG to introduce relevant clinical context generally improves performance, with the effect being more noticeable in the lower performing models. These findings suggest that reasoning LLMs are rapidly advancing toward becoming viable complementary decision-support tools in clinical practice. However, performance gaps persist, necessitating rigorous safeguards and continued human oversight—particularly in areas with identified limitations—to ensure patient safety.

## Materials and Methods

### Evaluation Datasets

For the evaluation we used two curated question banks from medical exam questionnaires with multiple-choice answers, one based on the MRCOG Part 2 examination and the other on the USMLE examination.

### MRCOG Part 2 Question Bank

We generated a question bank of 1392 questions MRCOG part 2 questions, consisting of 706 SBAs and 686 EMQs, taken from published books intended for revisions which are not believed to be part of the training data of the tested LLMs.

The pass mark for human expert candidates varies depending on the test while only about 20-30% of candidates pass^30^.

MRCOG Part 2 questions and answers were sourced from books with example questions used by medical students for revisions^30–35^. All selected questions were published after 2012 so they reflect the contemporary content of the examination.

Extraction of questions was performed using Claude 3.5, with each question, all possible answers, and the correct answer extracted into a CSV file for later testing. Each question was checked to be word-for-word identical to the original and LLM’s context was limited to the individual chapter/section of questions to minimise hallucination. For all questions the correct response was available from source, which served as a ground truth.

Questions were also collated and cleaned to be deduplicated across the different sources. The question prose was converted to lowercase alphanumeric characters, and manual cleaning of the questions was used to identify and resolve artifacts which were present in the source PDFs such as erroneous accented characters.

In this study, our reference to the copyrighted examination materials is minimal and serves strictly to evaluate Large Language Model performance, a purpose that aligns with fair dealing principles under UK copyright law (Copyright, Designs and Patents Act 1988). We do not reproduce the questions in their entirety, nor do we intend to substitute for or diminish the market of the original works.

### MedQA

The MedQA^3^ dataset is a multiple-choice question set derived from medical licensing exams in the United States (USMLE). The questions include vignette-style prompts (clinical scenarios) with four or five possible answers, and a single correct answer, closely resembling real medical board exam questions.

We generated a dataset of 945 MedQA questions, sourced from the repository github.com/jind11/MedQA, an open dataset of questions^3^ and filtered to those questions which reference women’s health terminology using a whitelist of terms and referring to a female patient scenario (See **Table S3** in Supplementary Tables for terms used). This ensured that women’s health questions only were included. All selected questions were further manually validated to ensure clinical relevance. The scope of questions is broader than the MRCOG Part 2, as the USMLE examination intends to test broad medical knowledge across specialties and not specialised Obs-Gynae knowledge. MedQA is not known to be used in training of medical LLMs, and it is being widely used for model evaluation ^36–39^; however, partial exposure of LLMs to some of the questions during training may have occurred.

Similarly to the MRCOG part 2 extraction, the MedQA question bank was generated by using GPT-4o for an initial extraction of questions, followed by manual review and cleaning of special characters. Each question was extracted alongside the possible answers/options and the correct answer that was used as ground truth.

### Model Variants

We evaluated a wide range of state of the art LLMs:

1. **General-Purpose LLMs** which are believed to be trained on a wide selection of text corpora from the internet. The specific models included in the evaluation are: OpenAI’s o1-preview, o3-mini, GPT-4o^4^, Meta’s Llama 3 (4 versions: 3.1-8b, 3.1-70b, 3.1-405b, 3.2-90b)^18^, Anthropic’s Claude (version: 3.5 Sonnet)^19^, Google’s Gemini (2 versions: 1.5 Flash, 1.5 Pro)^20^, Deepseek (version: R1-Reasoner)^21^
2. **Domain-Specialised Models**, which have been fine-tuned by including curated medical datasets in their training, in particular: MedLM^22^ which has been fine-tuned specifically for medical Q&A and MMed-Llama^23^ a model specifically trained to answer English medical questions.
3. **Augmented models using retrieval-augmented generation (RAG)** in which the question prompt was enriched with relevant context selected from embeddings generated from segments of 600 women’s health documents. We implemented RAG ‘on top’ of all the general-purpose models, to investigate if relevant context can improve their performance.

### Model naming & versions

Table 2 describes the exact model versions used and the naming we use to refer to models throughout the document. While we chose specific model releases for each model, proprietary models such as those coming from OpenAI, Anthropic & Gemini cannot be guaranteed to be stable over time as they are not released in an open-weight format.

**Table 2.** Model naming and versions key.

| Full Model Name | Publisher | Model Version | Simplified Name |
| --- | --- | --- | --- |
| Llama 3.1 405b | Meta | Llama-3.1-405B-Instruct | Llama 3.1-405b |
| Llama 3.2 90b | Meta | Llama-3.2-90b-vision-instruct | Llama 3.2-90b |
| Llama 3.1 70b | Meta | Llama-3.1-70B-Instruct | Llama 3.1-70b |
| Llama 3.1 405b | Meta | Llama-3.1-8B-Instruct | Llama 3.1-8b |
| R1 Reasoner | Deepseek | R1 Reasoner | DeepSeek R1 |
| o3-mini-high | OpenAI | o3-mini-2025-01-31,<br>set to “high” reasoning | o3-mini |
| o1-preview | OpenAI | o1-preview-2024-09-12 | o1-preview |
| GPT-4o | OpenAI | gpt-4o-2024-08-06 | GPT-4o |
| Claude 3.5 Sonnet | Anthropic | claude-3-5-sonnet 2024-06-20 | Claude 3.5 Sonnet |
| MedLM-Large | Google | medlm-large 2023-12-13 | MedLM |
| MMed-Llama-3 8b | <sup>23</sup> | MMed-Llama 3-8B-EnIns | MMed-Llama |
| Gemini 1.5 Pro | Google | gemini-1.5-pro-002 | Gemini 1.5 Pro |
| Gemini 1.5 Flash | Google | gemini-1.5-flash-002 | Gemini 1.5 Flash |

#### Prompt Engineering

For consistency we used the same simple prompt for every model, asking the model to give a single letter answer to a multiple choice question. In the RAG set up, the prompt was also consistent across all models, with wording added to consider additional medical context before presenting the question.

#### Evaluation Metrics

We report accuracy for each question bank separately as the percentage of correct answers over the entire question bank. To quantify uncertainty, we construct 95% confidence intervals using bootstrapping (see below).

**Retrieval-Augmented Generation Knowledge bank** with 600 documents manually selected from the MRCOG reading list^24^.

#### Retrieval-Augmented Generation

We take a standardised approach of: 1. Splitting documents into overlapping chunks in a recursive fashion 2. Tokenization and deduplication of chunks 3. Embedding chunks using a sentence transformer (text-embedding-ada-002) and storing them into a FAISS vector store to be used as a knowledge base 4. Setting up a cosine similarity distance metric for retrieval. During retrieval, 1. Each question used the same sentence transformer to create a question embedding 2. The question embedding is used for retrieval from the vector store using cosine similarity 3. The top 30 returned segments were reranked using the *colbert v2*.*0*^*40*^ reranker model to extract the 5 that contained the most clinically relevant information to the given query. 4. The final prompt is constructed to contain the text from the top 5 reranked segments, the question and the options.

#### Bootstrapping for Confidence intervals

All reported performance estimate and accompanying error bars utilised the same framework. A dataset was assembled with the complete list of questions, and the success/failure of the responses by model. This dataset was subset to the relevant model, categories, and question formats for the respective estimate, then was sampled using either a random non-repeating sample of size 50, or an ‘exam simulation’ sample. The exam simulation sample uses a random non-repeating sample of 50 SBAs and 50 EMQs.

5000 iterations of samples were taken. In the case of the exam simulations, the performance within a sample was a weighted average, with the % of correct SBAs contributing 40% and % of correct EMQs contributing 60% of the overall sample’s score. For the simple samples of size 50, a mean aggregation was used. The 2.5^th^ and 97.5^th^ percentile scores across the samples provided the lower and upper confidence intervals, and the mean across the samples provide the reported performance estimates.

## Author contributions

Conceptualization of the study was done by MI, NK, VO, TV, and CML. Data processing was carried out by VO and TV. NK and TV performed the coding and analysis. JS, RR, CG, CW, MG, NK, PS, IG, and CML were responsible for idea generation and discussion. The manuscript was written by MI, NK, VO, and TV. All authors contributed technical or methodological input to the study and provided substantial edits to the manuscript. All authors read and approved the final manuscript.

## Data availability

All data used in this study were obtained from publicly available sources. The specific sources for all datasets are cited in the reference list of this manuscript.

## Code availability

The code used in this study cannot be made publicly available due to intellectual property constraints. Additionally, the code includes proprietary methods developed specifically for the research. However, we will provide detailed descriptions of the methodologies and computational approaches upon request to the corresponding author.

