## Supplemental Tables for "Reasoning Over Pre-training: Evaluating LLM Performance and Augmentation in Women’s Health"

### Supplementary Tables

#### Table S1: Question type model performance detail

| Model | Question Type | Bootstrapped Accuracy Mean | CI Lower (2.5%) | CI Upper (97.5%) |
|---|---|---|---|---|
| Gemini 1.5 Pro | EMQ | 53.55% | 30% | 75% |
| Gemini 1.5 Pro | SBA | 59.42% | 40% | 80% |
| Gemini 1.5 Pro | MEDQA | 74.91% | 55% | 95% |
| Gemini 1.5 Flash | EMQ | 45.73% | 25% | 65% |
| Gemini 1.5 Flash | SBA | 50.68% | 30% | 70% |
| Gemini 1.5 Flash | MEDQA | 61.17% | 40% | 80% |
| Claude 3.5 Sonnet | EMQ | 57.98% | 35% | 80% |
| Claude 3.5 Sonnet | SBA | 61.53% | 40% | 80% |
| Claude 3.5 Sonnet | MEDQA | 78.65% | 60% | 95% |
| Claude 3.5 Sonnet RAG | EMQ | 59.49% | 40% | 80% |
| Claude 3.5 Sonnet RAG | SBA | 64.90% | 45% | 85% |
| Claude 3.5 Sonnet RAG | MEDQA | 73.13% | 55% | 90% |
| Deepseek R1 Reasoner | EMQ | 66.26% | 45% | 85% |
| Deepseek R1 Reasoner | SBA | 70.12% | 50% | 90% |
| Deepseek R1 Reasoner | MEDQA | 89.27% | 75% | 100% |
| Deepseek R1 Reasoner RAG | EMQ | 65.01% | 45% | 85% |
| Deepseek R1 Reasoner RAG | SBA | 70.70% | 50% | 90% |
| Deepseek R1 Reasoner RAG | MEDQA | 85.78% | 70% | 100% |
| MedLM MedLM | EMQ | 55.38% | 35% | 75% |
| MedLM MedLM | SBA | 54.38% | 30% | 75% |
| MedLM MedLM | MEDQA | 66.46% | 45% | 85% |
| OpenAI o3-mini-high RAG | EMQ | 67.18% | 45% | 85% |
| OpenAI o3-mini-high RAG | SBA | 71.27% | 50% | 90% |
| OpenAI o3-mini-high RAG | MEDQA | 92.90% | 80% | 100% |
| OpenAI o1-preview RAG | EMQ | 70.98% | 50% | 90% |
| OpenAI o1-preview RAG | SBA | 73.31% | 55% | 90% |
| OpenAI o1-preview RAG | MEDQA | 91.84% | 80% | 100% |
| OpenAI GPT4o RAG | EMQ | 51.62% | 30% | 70% |
| OpenAI GPT4o RAG | SBA | 62.61% | 40% | 80% |
| OpenAI GPT4o RAG | MEDQA | 83.18% | 65% | 100% |
| OpenAI GPT4o | EMQ | 33.89% | 15% | 55% |
| OpenAI GPT4o | SBA | 42.20% | 20% | 65% |
| OpenAI GPT4o | MEDQA | 86.25% | 70% | 100% |
| OpenAI o3-mini-high | EMQ | 65.84% | 45% | 85% |
| OpenAI o3-mini-high | SBA | 65.08% | 45% | 85% |
| OpenAI o3-mini-high | MEDQA | 93.04% | 80% | 100% |
| OpenAI o1-preview | EMQ | 69.06% | 50% | 90% |

| | | | | |
|---|---|---|---|---|
| OpenAI o1-preview | SBA | 71.42% | 50% | 90% |
| OpenAI o1-preview | MEDQA | 92.33% | 80% | 100% |
| Llama 3.1 70b | EMQ | 55.17% | 35% | 75% |
| Llama 3.1 70b | SBA | 59.71% | 40% | 80% |
| Llama 3.1 70b | MEDQA | 84.82% | 70% | 100% |
| Llama MMed-Llama-3-8B-enlns | EMQ | 35.14% | 15% | 55% |
| Llama MMed-Llama-3-8B-enlns | SBA | 45.93% | 25% | 70% |
| Llama MMed-Llama-3-8B-enlns | MEDQA | 78.96% | 60% | 95% |
| Llama 3.2 90b | EMQ | 54.56% | 35% | 75% |
| Llama 3.2 90b | SBA | 59.79% | 40% | 80% |
| Llama 3.2 90b | MEDQA | 84.66% | 70% | 100% |
| Llama 3.1 8b RAG | EMQ | 39.34% | 20% | 60% |
| Llama 3.1 8b RAG | SBA | 51.59% | 30% | 75% |
| Llama 3.1 8b RAG | MEDQA | 54.97% | 35% | 75% |
| Llama 3.1 8b | EMQ | 37.49% | 15% | 60% |
| Llama 3.1 8b | SBA | 47.24% | 25% | 70% |
| Llama 3.1 8b | MEDQA | 63.43% | 40% | 85% |
| Llama 3.1 405b | EMQ | 58.79% | 39.875% | 80% |
| Llama 3.1 405b | SBA | 63.61% | 40% | 85% |
| Llama 3.1 405b | MEDQA | 90.33% | 75% | 100% |
| Llama 3.1 405b RAG | EMQ | 61.73% | 40% | 85% |
| Llama 3.1 405b RAG | SBA | 65.55% | 45% | 85% |
| Llama 3.1 405b RAG | MEDQA | 82.74% | 65% | 100% |

### Table S2: Subdomain model performance detail

| Model | Subdomain | Bootstrapped Accuracy Mean | CI Lower (2.5%) | CI Upper (97.5%) |
|---|---|---|---|---|
| Gemini 1.5 Pro | Maternal Medicine and Medical Disorders in Pregnancy | 60.23% | 48% | 72% |
| Gemini 1.5 Pro | Fetal Medicine, Labour, Delivery and Postpartum Care | 47.78% | 36% | 60% |
| Gemini 1.5 Pro | Gynaecological Problems and Oncology | 62.14% | 50% | 74% |
| Gemini 1.5 Pro | Sexual and Reproductive Health | 60.98% | 52% | 70% |
| Gemini 1.5 Pro | Surgical Skills and Procedures | 45.42% | 38% | 54% |
| Gemini 1.5 Pro | Antenatal and Early Pregnancy Care | 55.33% | 42% | 68% |
| Gemini 1.5 Pro | Subfertility and Endocrinology | 50.77% | 40% | 62% |
| Gemini 1.5 Pro | Clinical Governance, Research and Teaching | 69.96% | 62% | 78% |
| Gemini 1.5 Flash | Antenatal and Early Pregnancy Care | 48.37% | 36% | 60% |
| Gemini 1.5 Flash | Fetal Medicine, Labour, Delivery and Postpartum Care | 39.47% | 28% | 52% |
| Gemini 1.5 Flash | Maternal Medicine and Medical Disorders in Pregnancy | 52.43% | 40% | 64% |
| Gemini 1.5 Flash | Gynaecological Problems and Oncology | 52.46% | 40% | 66% |
| Gemini 1.5 Flash | Sexual and Reproductive Health | 55.20% | 46% | 64% |
| Gemini 1.5 Flash | Subfertility and Endocrinology | 44.33% | 34% | 56% |
| Gemini 1.5 Flash | Surgical Skills and Procedures | 35.76% | 28% | 44% |
| Gemini 1.5 Flash | Clinical Governance, Research and Teaching | 64.06% | 56% | 74% |
| Claude 3.5 Sonnet | Sexual and Reproductive Health | 61.10% | 52% | 70% |
| Claude 3.5 Sonnet | Antenatal and Early Pregnancy Care | 61.61% | 50% | 74% |
| Claude 3.5 Sonnet | Maternal Medicine and Medical Disorders in Pregnancy | 62.60% | 52% | 74% |
| Claude 3.5 Sonnet | Fetal Medicine, Labour, Delivery and Postpartum Care | 52.88% | 40% | 66% |
| Claude 3.5 Sonnet | Gynaecological Problems and Oncology | 60.25% | 48% | 72% |
| Claude 3.5 Sonnet | Subfertility and Endocrinology | 61.81% | 52% | 72% |
| Claude 3.5 Sonnet | Surgical Skills and Procedures | 52.42% | 44% | 60% |
| Claude 3.5 Sonnet | Clinical Governance, Research and Teaching | 68.52% | 60% | 78% |
| Claude 3.5 Sonnet RAG | Antenatal and Early Pregnancy Care | 63.54% | 52% | 76% |
| Claude 3.5 Sonnet RAG | Fetal Medicine, Labour, Delivery and Postpartum Care | 59.15% | 48% | 70% |
| Claude 3.5 Sonnet RAG | Maternal Medicine and Medical Disorders in Pregnancy | 61.22% | 50% | 72% |
| Claude 3.5 Sonnet RAG | Gynaecological Problems and Oncology | 64.94% | 54% | 76% |
| Claude 3.5 Sonnet RAG | Sexual and Reproductive Health | 59.90% | 52% | 68% |
| Claude 3.5 Sonnet RAG | Subfertility and Endocrinology | 58.61% | 48% | 68% |
| Claude 3.5 Sonnet RAG | Surgical Skills and Procedures | 61.77% | 54% | 70% |

| | | | | |
|---|---|---|---|---|
| Claude 3.5 Sonnet RAG | Clinical Governance, Research and Teaching | 68.70% | 60% | 78% |
| Deepseek R1 Reasoner | Antenatal and Early Pregnancy Care | 67.72% | 56% | 78% |
| Deepseek R1 Reasoner | Fetal Medicine, Labour, Delivery and Postpartum Care | 62.59% | 50% | 74% |
| Deepseek R1 Reasoner | Maternal Medicine and Medical Disorders in Pregnancy | 70.10% | 60% | 80% |
| Deepseek R1 Reasoner | Gynaecological Problems and Oncology | 71.88% | 60% | 84% |
| Deepseek R1 Reasoner | Sexual and Reproductive Health | 59.80% | 50% | 68% |
| Deepseek R1 Reasoner | Subfertility and Endocrinology | 66.79% | 56% | 78% |
| Deepseek R1 Reasoner | Surgical Skills and Procedures | 66.82% | 60% | 74% |
| Deepseek R1 Reasoner | Clinical Governance, Research and Teaching | 73.05% | 64% | 82% |
| Deepseek R1 Reasoner RAG | Sexual and Reproductive Health | 58.72% | 50% | 68% |
| Deepseek R1 Reasoner RAG | Clinical Governance, Research and Teaching | 75.09% | 68% | 82% |
| Deepseek R1 Reasoner RAG | Gynaecological Problems and Oncology | 72.58% | 62% | 84% |
| Deepseek R1 Reasoner RAG | Fetal Medicine, Labour, Delivery and Postpartum Care | 62.30% | 50% | 74% |
| Deepseek R1 Reasoner RAG | Surgical Skills and Procedures | 75.33% | 68% | 82% |
| Deepseek R1 Reasoner RAG | Antenatal and Early Pregnancy Care | 67.34% | 56% | 78% |
| Deepseek R1 Reasoner RAG | Maternal Medicine and Medical Disorders in Pregnancy | 67.25% | 56% | 78% |
| Deepseek R1 Reasoner RAG | Subfertility and Endocrinology | 64.80% | 54% | 74% |
| MedLM MedLM | Maternal Medicine and Medical Disorders in Pregnancy | 58.47% | 46% | 70% |
| MedLM MedLM | Fetal Medicine, Labour, Delivery and Postpartum Care | 46.12% | 34% | 58% |
| MedLM MedLM | Gynaecological Problems and Oncology | 58.03% | 46% | 70% |
| MedLM MedLM | Sexual and Reproductive Health | 62.12% | 52% | 70% |
| MedLM MedLM | Surgical Skills and Procedures | 43.15% | 36% | 52% |
| MedLM MedLM | Antenatal and Early Pregnancy Care | 52.97% | 40% | 66% |
| MedLM MedLM | Subfertility and Endocrinology | 57.12% | 46% | 68% |
| MedLM MedLM | Clinical Governance, Research and Teaching | 64.66% | 56% | 74% |
| OpenAI o3-mini-high RAG | Sexual and Reproductive Health | 66.83% | 58% | 76% |
| OpenAI o3-mini-high RAG | Clinical Governance, Research and Teaching | 77.16% | 70% | 84% |
| OpenAI o3-mini-high RAG | Gynaecological Problems and Oncology | 70.80% | 60% | 82% |

| | | | | |
|---|---|---|---|---|
| OpenAI o3-mini-high RAG | Fetal Medicine, Labour, Delivery and Postpartum Care | 66.09% | 54% | 78% |
| OpenAI o3-mini-high RAG | Surgical Skills and Procedures | 74.84% | 68% | 82% |
| OpenAI o3-mini-high RAG | Antenatal and Early Pregnancy Care | 67.74% | 56% | 80% |
| OpenAI o3-mini-high RAG | Maternal Medicine and Medical Disorders in Pregnancy | 72.22% | 62% | 82% |
| OpenAI o3-mini-high RAG | Subfertility and Endocrinology | 62.73% | 52% | 74% |
| OpenAI o1-preview RAG | Sexual and Reproductive Health | 69.20% | 62% | 78% |
| OpenAI o1-preview RAG | Antenatal and Early Pregnancy Care | 70.80% | 60% | 82% |
| OpenAI o1-preview RAG | Maternal Medicine and Medical Disorders in Pregnancy | 74.06% | 64% | 84% |
| OpenAI o1-preview RAG | Fetal Medicine, Labour, Delivery and Postpartum Care | 66.31% | 54% | 78% |
| OpenAI o1-preview RAG | Gynaecological Problems and Oncology | 74.71% | 64% | 86% |
| OpenAI o1-preview RAG | Subfertility and Endocrinology | 69.69% | 60% | 80% |
| OpenAI o1-preview RAG | Surgical Skills and Procedures | 79.54% | 72% | 86% |
| OpenAI o1-preview RAG | Clinical Governance, Research and Teaching | 74.71% | 66% | 82% |
| OpenAI GPT4o RAG | Sexual and Reproductive Health | 58.82% | 50% | 68% |
| OpenAI GPT4o RAG | Antenatal and Early Pregnancy Care | 58.51% | 46% | 70% |
| OpenAI GPT4o RAG | Maternal Medicine and Medical Disorders in Pregnancy | 65.44% | 54% | 76% |
| OpenAI GPT4o RAG | Fetal Medicine, Labour, Delivery and Postpartum Care | 55.17% | 44% | 68% |
| OpenAI GPT4o RAG | Gynaecological Problems and Oncology | 54.79% | 42% | 68% |
| OpenAI GPT4o RAG | Subfertility and Endocrinology | 51.24% | 40% | 62% |
| OpenAI GPT4o RAG | Surgical Skills and Procedures | 56.47% | 48% | 64% |
| OpenAI GPT4o RAG | Clinical Governance, Research and Teaching | 51.30% | 42% | 60% |
| OpenAI GPT4o | Maternal Medicine and Medical Disorders in Pregnancy | 44.71% | 34% | 56% |
| OpenAI GPT4o | Fetal Medicine, Labour, Delivery and Postpartum Care | 35.95% | 24% | 48% |
| OpenAI GPT4o | Gynaecological Problems and Oncology | 32.47% | 20% | 44% |
| OpenAI GPT4o | Sexual and Reproductive Health | 42.51% | 34% | 50% |
| OpenAI GPT4o | Surgical Skills and Procedures | 27.48% | 20% | 34% |
| OpenAI GPT4o | Antenatal and Early Pregnancy Care | 44.73% | 32% | 56% |
| OpenAI GPT4o | Subfertility and Endocrinology | 37.55% | 28% | 48% |
| OpenAI GPT4o | Clinical Governance, Research and Teaching | 37.16% | 28% | 46% |
| OpenAI o3-mini-high | Maternal Medicine and Medical Disorders in Pregnancy | 77.53% | 68% | 88% |

| | | | | |
|---|---|---|---|---|
| OpenAI o3-mini-high | Fetal Medicine, Labour, Delivery and Postpartum Care | 60.20% | 48% | 72% |
| OpenAI o3-mini-high | Gynaecological Problems and Oncology | 68.55% | 56% | 80% |
| OpenAI o3-mini-high | Sexual and Reproductive Health | 62.54% | 54% | 72% |
| OpenAI o3-mini-high | Surgical Skills and Procedures | 50.64% | 42% | 58% |
| OpenAI o3-mini-high | Antenatal and Early Pregnancy Care | 62.40% | 50% | 74% |
| OpenAI o3-mini-high | Subfertility and Endocrinology | 64.26% | 54% | 74% |
| OpenAI o3-mini-high | Clinical Governance, Research and Teaching | 72.76% | 64% | 82% |
| OpenAI o1-preview | Sexual and Reproductive Health | 70.49% | 62% | 78% |
| OpenAI o1-preview | Antenatal and Early Pregnancy Care | 69.55% | 58% | 80% |
| OpenAI o1-preview | Maternal Medicine and Medical Disorders in Pregnancy | 72.36% | 62% | 82% |
| OpenAI o1-preview | Fetal Medicine, Labour, Delivery and Postpartum Care | 63.58% | 52% | 76% |
| OpenAI o1-preview | Gynaecological Problems and Oncology | 74.79% | 64% | 86% |
| OpenAI o1-preview | Subfertility and Endocrinology | 68.65% | 58% | 78% |
| OpenAI o1-preview | Surgical Skills and Procedures | 70.15% | 62% | 78% |
| OpenAI o1-preview | Clinical Governance, Research and Teaching | 71.86% | 64% | 80% |
| Llama 3.1 70b | Sexual and Reproductive Health | 57.72% | 48% | 66% |
| Llama 3.1 70b | Clinical Governance, Research and Teaching | 66.13% | 58% | 74% |
| Llama 3.1 70b | Gynaecological Problems and Oncology | 62.69% | 50% | 74% |
| Llama 3.1 70b | Fetal Medicine, Labour, Delivery and Postpartum Care | 48.99% | 36% | 62% |
| Llama 3.1 70b | Surgical Skills and Procedures | 47.75% | 40% | 56% |
| Llama 3.1 70b | Antenatal and Early Pregnancy Care | 56.13% | 44% | 68% |
| Llama 3.1 70b | Maternal Medicine and Medical Disorders in Pregnancy | 63.78% | 52% | 76% |
| Llama 3.1 70b | Subfertility and Endocrinology | 54.72% | 44% | 66% |
| Llama MMed-Llama-3-8B-enlns | Sexual and Reproductive Health | 39.02% | 30% | 48% |
| Llama MMed-Llama-3-8B-enlns | Antenatal and Early Pregnancy Care | 37.78% | 26% | 50% |
| Llama MMed-Llama-3-8B-enlns | Maternal Medicine and Medical Disorders in Pregnancy | 44.17% | 32% | 56% |
| Llama MMed-Llama-3-8B-enlns | Fetal Medicine, Labour, Delivery and Postpartum Care | 38.44% | 26% | 50% |
| Llama MMed-Llama-3-8B-enlns | Gynaecological Problems and Oncology | 44.78% | 32% | 56% |
| Llama MMed-Llama-3-8B-enlns | Subfertility and Endocrinology | 33.19% | 24% | 44% |
| Llama MMed-Llama-3-8B-enlns | Surgical Skills and Procedures | 35.75% | 28% | 44% |

| | | | | |
|---|---|---|---|---|
| Llama MMed-Llama-3-8B-enlns | Clinical Governance, Research and Teaching | 53.59% | 44% | 64% |
| Llama 3.2 90b | Antenatal and Early Pregnancy Care | 54.52% | 42% | 66% |
| Llama 3.2 90b | Fetal Medicine, Labour, Delivery and Postpartum Care | 48.92% | 36% | 62% |
| Llama 3.2 90b | Maternal Medicine and Medical Disorders in Pregnancy | 62.90% | 52% | 74% |
| Llama 3.2 90b | Gynaecological Problems and Oncology | 62.23% | 50% | 74% |
| Llama 3.2 90b | Sexual and Reproductive Health | 58.86% | 50% | 68% |
| Llama 3.2 90b | Subfertility and Endocrinology | 54.61% | 44% | 66% |
| Llama 3.2 90b | Surgical Skills and Procedures | 47.77% | 40% | 56% |
| Llama 3.2 90b | Clinical Governance, Research and Teaching | 67.02% | 58% | 76% |
| Llama 3.1 8b RAG | Antenatal and Early Pregnancy Care | 46.14% | 34% | 58% |
| Llama 3.1 8b RAG | Fetal Medicine, Labour, Delivery and Postpartum Care | 42.72% | 30% | 56% |
| Llama 3.1 8b RAG | Maternal Medicine and Medical Disorders in Pregnancy | 46.11% | 34% | 58% |
| Llama 3.1 8b RAG | Gynaecological Problems and Oncology | 48.59% | 36% | 62% |
| Llama 3.1 8b RAG | Sexual and Reproductive Health | 40.39% | 32% | 50% |
| Llama 3.1 8b RAG | Subfertility and Endocrinology | 36.80% | 26% | 48% |
| Llama 3.1 8b RAG | Surgical Skills and Procedures | 47.49% | 40% | 56% |
| Llama 3.1 8b RAG | Clinical Governance, Research and Teaching | 56.38% | 48% | 66% |
| Llama 3.1 8b | Maternal Medicine and Medical Disorders in Pregnancy | 42.96% | 32% | 54% |
| Llama 3.1 8b | Fetal Medicine, Labour, Delivery and Postpartum Care | 37.61% | 26% | 50% |
| Llama 3.1 8b | Gynaecological Problems and Oncology | 47.04% | 34% | 60% |
| Llama 3.1 8b | Sexual and Reproductive Health | 39.26% | 30% | 48% |
| Llama 3.1 8b | Surgical Skills and Procedures | 32.34% | 24% | 40% |
| Llama 3.1 8b | Antenatal and Early Pregnancy Care | 40.23% | 28% | 52% |
| Llama 3.1 8b | Subfertility and Endocrinology | 42.24% | 32% | 54% |
| Llama 3.1 8b | Clinical Governance, Research and Teaching | 54.26% | 44% | 64% |
| Llama 3.1 405b | Sexual and Reproductive Health | 58.79% | 50% | 68% |
| Llama 3.1 405b | Antenatal and Early Pregnancy Care | 57.02% | 46% | 70% |
| Llama 3.1 405b | Maternal Medicine and Medical Disorders in Pregnancy | 67.35% | 56% | 78% |
| Llama 3.1 405b | Fetal Medicine, Labour, Delivery and Postpartum Care | 55.12% | 42% | 68% |
| Llama 3.1 405b | Gynaecological Problems and Oncology | 65.23% | 54% | 76% |
| Llama 3.1 405b | Subfertility and Endocrinology | 60.07% | 50% | 70% |
| Llama 3.1 405b | Surgical Skills and Procedures | 53.07% | 46% | 62% |

| | | | | |
|---|---|---|---|---|
| Llama 3.1 405b | Clinical Governance, Research and Teaching | 72.74% | 64% | 80% |
| Llama 3.1 405b RAG | Antenatal and Early Pregnancy Care | 61.29% | 50% | 72% |
| Llama 3.1 405b RAG | Fetal Medicine, Labour, Delivery and Postpartum Care | 59.36% | 48% | 70% |
| Llama 3.1 405b RAG | Maternal Medicine and Medical Disorders in Pregnancy | 69.69% | 58% | 80% |
| Llama 3.1 405b RAG | Gynaecological Problems and Oncology | 65.25% | 52% | 76% |
| Llama 3.1 405b RAG | Sexual and Reproductive Health | 56.54% | 48% | 66% |
| Llama 3.1 405b RAG | Subfertility and Endocrinology | 59.31% | 48% | 70% |
| Llama 3.1 405b RAG | Surgical Skills and Procedures | 66.21% | 58% | 74% |
| Llama 3.1 405b RAG | Clinical Governance, Research and Teaching | 69.59% | 62% | 78% |

### Table S3: Whitelisted terms used for MEDQA question selection

| Term |
|---|
| Gestational |
| Pre-eclampsia |
| Eclampsia |
| Fetal monitoring |
| Placenta previa |
| Placenta |
| Amniocentesis |
| Chorionic villus sampling |
| Fetal heart rate |
| Labor dystocia |
| Cervical effacement |
| Cervical dilation |
| Cervical |
| Episiotomy |
| Postpartum hemorrhage |
| Postpartum |
| Lochia |
| Preeclampsia |
| Oligohydramnios |
| Polyhydramnios |
| Fetal macrosomia |
| Intrauterine growth restriction |
| IUGR |
| Teratogen |
| Ectopic pregnancy |
| Gestational diabetes |
| In vitro fertilization |
| IVF |
| Intrauterine insemination |
| IUI |
| Anovulation |
| Polycystic ovary syndrome |
| PCOS |
| Endometriosis |
| Hysterosalpingography |
| Laparoscopy |
| Menorrhagia |

| |
|---|
| Dysmenorrhea |
| Amenorrhea |
| Oophorectomy |
| Salpingectomy |
| Hysterectomy |
| Myomectomy |
| Adenomyosis |
| Bartholin cyst |
| Cervical dysplasia |
| HPV vaccination |
| Pap smear |
| smear |
| Colposcopy |
| LEEP procedure |
| LEEP |
| Vaginal atrophy |
| Hormone replacement therapy |
| HRT |
| Menopause |
| Menopausal |
| Premenopausal |
| Postmenopausal |
| Osteoporosis |
| Breastfeeding |
| Lactation consultant |
| Ovary |
| Fallopian tube |
| Uterus |
| Endometrium |
| Myometrium |
| Perimetrium |
| Cervix |
| Vagina |
| Vaginal |
| Vulva |
| Clitoris |
| Labia majora |
